# Dynamics of within-host *Mycobacterium tuberculosis* diversity and heteroresistance during treatment

**DOI:** 10.1101/2020.02.03.20019786

**Authors:** Camus Nimmo, Kayleen Brien, James Millard, Alison D. Grant, Nesri Padayatchi, Alexander S. Pym, Max O’Donnell, Richard Goldstein, Judith Breuer, François Balloux

## Abstract

**Background:** Studying within-host genetic diversity of *Mycobacterium tuberculosis* (*Mtb*) in patients during treatment may identify adaptations to antibiotic and immune pressure. Understanding the significance of genetic heteroresistance, and more specifically heterozygous resistance-associated variants (RAVs), is clinically important given increasing use of rapid molecular tests and whole genome sequencing (WGS).

**Methods:** We analyse data from six studies in KwaZulu-Natal, South Africa. Most patients (>75%) had baseline rifampicin-resistance. Sputum was collected for culture at baseline and at between two and nine intervals until month six. Positive cultures underwent WGS. Mixed infections and reinfections were excluded from analysis.

**Findings:** Baseline *Mtb* overall genetic diversity (at treatment initiation or major change to regimen) was associated with cavitary disease, not taking antiretroviral therapy if HIV infected, infection with lineage 2 strains and absence of second-line drug resistance on univariate analyses. Baseline genetic diversity was not associated with six-month outcome. Genetic diversity increased from baseline to weeks one and two before returning to previous levels. Baseline genetic heteroresistance was most common for bedaquiline (6/10 [60%] of isolates with RAVs) and fluoroquinolones (9/62 [13%]). Most patients with heterozygous RAVs on WGS with sequential isolates available demonstrated persistence or fixation (17/20, 85%). New RAVs emerged in 9/286 (3%) patients during treatment. We could detect low-frequency RAVs preceding emergent resistance in only one case, although validation of deep sequencing to detect rare variants is required.

**Interpretation:** In this study of single-strain *Mtb* infections, baseline within-host bacterial genetic diversity did not predict outcome but may reveal adaptations to host and drug pressures. Predicting emergent resistance from low-frequency RAVs requires further work to separate transient from consequential mutations.

**Funding:** Wellcome Trust, NIH/NIAID

## Introduction

The *Mycobacterium tuberculosis* complex (MTBC), consisting of seven human-adapted lineages as well as several animal-adapted strains which may also cause human disease, is a major human pathogen. The most prominent species in the complex, *Mycobacterium tuberculosis* (*Mtb*), represents the infectious agent causing most deaths worldwide(1). *Mtb* has traditionally been viewed as a genetically homogenous bacterium that has evolved into a specialised human pathogen with a lower mutation rate than most other bacteria and no accessory genome or potential for horizontal gene transfer(2). More recent work has identified significant within-host *Mtb* genetic diversity which may result from mixed infection with multiple strains or within-host microevolution of a single infecting strain(3). Such genetic diversity is not surprising given that *Mtb* infections typically last months to years and within-host bacterial populations may peak at over one billion colony forming units. At one extreme, up to 50 consensus-level SNP differences having been reported to occur over the duration of infection in patients with advanced disease when sampling from multiple body sites(4). While genetic diversity arising from mixed infections has been associated with poor clinical outcomes(5,6), the effect of within-host diversity of single strain infections is undefined.

Within-host genetic diversity reflects the extent to which genomes (in this case *Mtb* genomes originating from individual bacteria) within a population (comprising all bacteria in the cultured isolate) are genetically distinct. It can be measured by nucleotide diversity (π), which is the mean number of nucleotide substitutions per site between any two randomly selected DNA sequences in a population, and takes a value between 0 and 1(7). We refer to any site carrying more than one allele as ‘heterozygous’, following standard practice in the population genetics literature to define multi-allelic loci, irrespective of ploidy(8–12).

Understanding factors affecting within-host *Mtb* genetic diversity may offer insights into mechanisms controlling bacterial replication and evolution. Studying sequential isolates from individual patients over the course of treatment could help identify signatures of bacterial adaptation to drug treatment and the host immune environment. Previous studies of within-host diversity have focused on reanalysis of published sequences from patients who have failed treatment. One detailed study of five patients revealed that overall *Mtb* genomic diversity increased with disease severity and was particularly high in pre-mortem isolates from two patients presumably due to high bacterial load(13). The most diverse genes were those involved in production of cell envelope lipids. No evidence for a decrease in diversity during treatment or any effect of *Mtb* lineage or drug resistance profile was found, and HIV statuses were not available for analysis. A larger analysis of combined data from eight publicly available studies reporting patients who failed treatment found that genes associated with antibiotic resistance displayed highest diversity, while the within-host diversity across remaining gene classes (essential, non-essential, PE/PPE genes and antigen genes) seemed unaffected(14).

However, when reanalysing data collected by other groups, it may be difficult to account for the role that pathogen culture and subsequent subculturing steps may have on genetic diversity through random loss or selection of culture-adapted subpopulations. We and others have shown that culture-independent sequencing of *Mtb* directly from sputum identifies more genetic diversity than sequencing from culture(15,16), while other studies have shown subculture can lead to loss of heterozygous resistance-associated variants (RAVs)(17). Furthermore, detailed analysis of low-frequency variants can be affected by errors induced during PCR steps of genomic library preparation(18) and incorrect mapping of contaminant DNA(19).

Mixed populations of wild-type alleles and RAVs confers heteroresistance, where populations of resistant and susceptible bacteria co-exist within the same host. This may occur as the result of differential drug penetration to spatially and pathologically distinct lung regions(20) leading in effect to monotherapy and subsequent resistance acquisition or survival of susceptible bacteria. Such mixed populations of wild type alleles and RAVs have been termed macroheteroresistance when RAVs are present at ≥5% frequency, a threshold above which they would be identified by most standard WGS pipelines, and microheteroresistance when RAVs are at <5% frequency(21). Several case reports have identified heterozygous RAVs (hetRAVs) that have increased in frequency over the course of treatment(22–24) leading to fixed resistance, including variants originally identified at <1% frequency(25), and genetic microheteroresistance has been identified predating acquired phenotypic resistance(26). However, due to high levels of turnover of low-frequency variants, it may be difficult to predict which hetRAVs are likely to persist or become fixed and which ones will disappear. HetRAVs are likely to be variably identified by current diagnostics(27,28) and so it may be difficult to predict whether such resistance is taken into account when clinical drug treatment regimens were designed for these patients.

The limit of detection for heterozygous variants depends on several factors including the number of reads covering a site, the minimum number of supporting reads required to call a variant and error rates at each step of the sequencing pipeline. For example, in this study where we have included sequences with a minimum 55x mean genome coverage with four reads required to call a variant, the limit of detection would be an allele frequency of 7%. We also use deep sequencing to >3000x coverage, where using a minimum of 6 supporting reads gives resolution to identify variants down to 0.2% frequency. However, the important caveat to this is that as coverage depth increases and the limit of detection decreases, the potential false positive rate due to PCR or sequencing errors increases. Using a base quality score of >30 (an error of 1 in 1000) and published DNA polymerase error rates(29), a base error rate of 0.1-0.2% would be expected, although rates up to 3% have been previously reported in some genomic regions(30).

In this study we used prospectively enrolled patients with TB to identify factors associated with within-host *Mtb* genetic diversity in single strain infections and assess whether this type of genetic diversity is associated with poor outcomes. We then investigated the outcomes of genetic heteroresistance and whether presence of microheteroresistance could predict the emergence of macroheteroresistance.

## Methods

### Patient recruitment

We analysed data from five observational cohort studies and a randomised controlled trial in KwaZulu-Natal, South Africa, as described in Table 1, conducted between 2016 and 2019. We selected mostly cohorts that recruited patients with DR-TB, as these patients have previously experienced the worst outcomes and most problems with acquired resistance, as well as one drug-susceptible cohort. Patients were recruited before or on the day of initiating treatment or undergoing a major change in treatment due to a change in drug resistance profile or clinical treatment failure. To be classified a major change in treatment regimen, the patient had to be starting at least two new drugs, be culture positive and have a previous regimen that was likely to be ineffective based on drug susceptibility testing (DST) results. At enrolment, all patients completed a study questionnaire and provided a sputum sample. Patients were reassessed at the intervals described in Table 1 with a follow-up questionnaire and repeat sputum sample (except in Hlabisa DR-TB cohort where follow-up involved completion of clinical record forms only). All patients provided written informed consent to participate. All study protocols were approved by the University of KwaZulu-Natal Biomedical Research Ethics Committee. PRAXIS was also approved by Columbia University Review Board and the Hlabisa DR-TB study by the London School of Hygiene & Tropical Medicine Ethics Committee.

**Table 1.** Description of cohort studies from which patients in this analysis were included. BL = baseline, W1 = week 1 etc, M1 = month 1 etc, EndTX = end of treatment, MDR = multidrug-resistant TB, XDR = extensively drug-resistant TB. Isolates with minimum 55x mean genome coverage were included in the analysis of heteroresistance and sequential genetic diversity. Isolates with minimum 80x mean genome coverage were included in the analysis of baseline genetic diversity. Number of patients with a baseline isolate are shown, with the number of those who had at least one sequential isolate shown in brackets. ClinicalTrials.gov registration number for PRAXIS study shown.

| Study number | Study name | Design | Patients recruited | Sputum collection timepoints | Patients with baseline (and ≥1 sequential) isolates by minimum coverage |  |
| --- | --- | --- | --- | --- | --- | --- |
|  |  |  |  |  | 55x | 80x |
| **1** | PRAXIS Aim 1 (NCT03162107) | Observational cohort study | MDR / XDR Starting bedaquiline | BL, M1, M2, M3, M4, M5, M6, EndTX | 57 (23) | 49 |
| **2** | PRAXIS Aim 2 (NCT03162107) | Randomised controlled trial | MDR / XDR Starting bedaquiline | BL, M2, M6, EndTX | 35 (6) | 38 |
| **3** | CUBS (PZAP arm) | Observational cohort study | MDR Treated with pyrazinamide | BL, W1, W2, W4, W6, M2, M3, M4, M5, M6 | 79 (54) | 72 |
| **4** | REPORT-SA | Observational cohort study | DS | BL, W1, W3, M2, M6 (EndTX) | 66 (33) | 67 |
| **5** | Hlabisa DR-TB | Observational cohort study | MDR | BL | 113 (N/A) | 108 |
| **6** | CUBS (FIND arm) | Observational cohort study | MDR | BL, M2, M6, EndTX | 49 (2) | 18 |
|  | Total |  |  |  | 399<br>(118) | 352 |

Every patient from these cohorts was included if they were enrolled prior to February 2019 and were culture positive with a baseline *Mtb* isolate with sufficient mean genome-wide coverage depth for the proposed analysis: 80x for evaluation of baseline diversity, 55x for evaluation of diversity and heteroresistance in sequential isolates. Minimum coverage thresholds were chosen to maximise sequencing coverage depth, and therefore ability to identify low level variation, while ensuring that sufficient numbers of samples fulfilled the criteria for inclusion. A lower threshold was used for the sequential analysis as some follow-up samples did not meet the minimum 80x coverage depth threshold. As described below, sequences for diversity analyses were downsampled to a standard mean coverage, while original coverage was used for heteroresistance. A list of isolates included in each analysis and associated accession numbers is available in the Supplementary Spreadsheet.

### Microbiology

All sputum samples were decontaminated, homogenised and inoculated into liquid mycobacterial growth indicator tube (MGIT) and on Middlebrook 7H11 solid agar for *Mtb* culture. Positive cultures underwent phenotypic DST for first- and second-line drugs on 7H11 agar using the 1% agar proportion method(31). The critical concentrations used are listed in the Supplementary Spreadsheet.

### Genome Sequencing

DNA was extracted for whole genome sequencing (WGS) from all positive MGIT cultures by mechanical ribolysis before purification with AMPure XP beads (Beckman Coulter, IN, USA) (full description in Supplementary Methods 2). When MGIT cultures yielded insufficient DNA for sequencing or were contaminated, DNA extraction from the parallel 7H11 culture or a 7H9 subculture was attempted. Given that multiple subculturing steps have been shown to reduce diversity, at most a single subculture step was used if required. Sequencing libraries were prepared with NEBNext Ultra II DNA (New England Biolabs, MA, USA) according to the manufacturer’s instructions, with between four and fourteen PCR cycles depending on input DNA concentration. Batches of 48 multiplexed isolates were sequenced on a NextSeq 500 (Illumina, CA, USA) with 300-cycle paired end runs using a mid-output kit.

Where emergent RAVs were identified by WGS, targeted deep sequencing was performed retrospectively on stored surplus extracted DNA from preceding isolates. A custom-designed SureSelect bait set (Agilent) covering genes associated with drug resistance comprising approximately 3% of the genome was designed and used for target capture of *Mtb* DNA from drug resistance-associated genes (Supplementary Table 1). Overlapping 120 base pair RNA baits were designed using SureDesign (Agilent) to cover the entire positive strand of over 800 *Mtb* genomes downloaded from NCBI Sequencing Read Archive selected to cover the global genetic diversity of *Mtb*. Bait design is freely available from the authors on request. Genomic DNA was fragmented to approximately 330 base pairs, which is longer than the standard sequencing read of 150 base pairs. Hybridisation and library preparation were performed with the SureSelectXT Low Input kit (Agilent) protocol according to the manufacturer’s instructions, including 20-23 PCR cycles depending on input concentration. Genomic libraries were multiplexed with 96 isolates sequenced on a NextSeq 500 (Illumina, CA, USA) 300-cycle paired end run using a mid-output kit.

### Bioinformatic Analysis

Raw sequencing reads had adapters removed with Trim Galore v0.3.7(32) and were mapped to the H37Rv reference genome (NC_000962.3) with BBMap v38.32(33) with a 98% identity threshold. Mapped reads were sorted and de-duplicated with Picard Tools v2.20(34) and mean genome coverage depth was checked with Qualimap v2.21(35). FreeBayes v1.3.1(36) was used to call variants with a specified minimum mapping quality 20 and base quality 30. For a variant to be called, ≥4 supporting reads were required for WGS and ≥6 supporting reads for targeted deep sequencing, including at least one on each strand with coverage ≥10. Variants were classed as heterozygous if present at <95% frequency or fixed if present at ≥95% frequency. Variants falling within repetitive regions poorly resolved by short read mapping (PE/PPE and *esx* genes and those categorised as insertion sequences and phages(37)) were excluded from the analysis.

Two steps were used in order to identify mixed infections. First, isolates with evidence of heterozygous SNPs at established lineage-specific sites(38) were classified as mixed infections. Then, to identify mixed infections of strains from the same sublineage (which would share sublineage-specific SNPs), we used a previously described method which identifies isolates with high numbers of heterozygous SNPs and then measures the degree of allele frequency clustering to discriminate within-host evolution of a single strain from infection with two genetically distinct strains(9).

The level of isolate contamination was estimated using Kraken2(39) to classify the proportion of *k*-mers allocated to the genus *Mycobacterium* using the MiniKraken2 v2 database (updated 23/04/2019). To prevent differences in coverage depth potentially affecting measures of π, aligned sequences were downsampled using Picard Tools v2.20(34) DownSampleSam. For baseline analysis, sequences were downsampled to mean 80x coverage depth and for sequential analysis to mean 55x coverage. Sequences were not downsampled for the analysis of heteroresistance. Within-isolate π was calculated by an in-house program that has been described previously(40) using base counts extracted from mapped alignments by bam-readcount(41) with mapping quality >20 and base quality >30. Diversity of individual genes was calculated using Popoolation v1.2(42) with the following parameters: pool size 500, minimum coverage 4, minimum count 2, minimum quality 30. For the comparison of diversity at the gene level, coding genes were annotated with one of seven functional categories as described in Mycobrowser(37). For comparability, we also labelled each gene as antibiotic resistance (*Rv0678, Rv1979c* and *rplC* were recategorised as antibiotic resistance genes), antigen, essential or non-essential as described previously(14) (PE/PPE genes were excluded from our analysis).

We first aimed to determine if the number of PCR cycles in library preparation or proportion of reads assigned to *Mycobacteria* in the isolate affected within-patient genetic diversity, as each PCR cycle comes with a theoretical risk of introducing base errors. We also investigated the possibility that contaminating reads erroneously mapped to *Mycobacteria* may inflate the genetic diversity.

For identification of RAVs, we used a curated list of resistance-conferring mutations adapted from published sources(43,44) as listed in Supplementary Methods 3. We supplemented these mutations lists with more recent published data for new and repurposed drugs(45–48). For mutations in the bedaquiline/clofazimine resistance-associated gene *Rv0678* and delamanid resistance-associated genes *fbiA*/*B*/*C, fgd1* and *ddn*, where a wide array of different mutations leading to loss of function occur, we took a more sensitive approach and manually evaluated all non-synonymous SNPs and indels for potential to cause resistance. We assumed that all loss of function mutations (frameshift or premature stop codons) to be potentially causative of resistance, as well as any SNPs that had previously been reported to associated with resistance.

### Clinical Assessment

Structured clinical record forms were completed at all visits by dedicated research nurses. Responses were taken from patients verbally as well as from clinic/hospital records. Chest radiography findings were recorded as documented by the treating clinician. Six-month outcome was assessed as favourable (alive, on treatment, *Mtb* culture negative at months 5 and 6) or unfavourable (death, loss to follow-up, culture positive at month 5 or 6).

### Statistical analysis

Statistical analyses were performed using Stata/IC v15.1 (StataCorp, TX, USA). Distributions of non-parametric data were compared with Wilcoxon rank-sum test. Binary outcomes were assessed by logistic regression and continuous variables with linear regression. The Benjamini-Hochberg procedure was used to control for multiple tests when evaluating change in median π between baseline and sequential timepoints with Q (false positive rate) of 0.1, which should yield less than one false positive test when performing seven tests.

## Results

### Patient selection and identification of mixed infection

A total of 399 from the 710 patients recruited into six different studies described in Table 1 were included in this study. The most common reason for exclusion were lack of a positive *Mtb* culture for sequencing. Treatment data were available for 334/399 (84%) patients and are described in the next paragraph. In some of these 334 cases there was discordance between the initial clinical resistance profile upon which drug treatment decisions were based and the study WGS profile: 2/66 (3%) patients initially diagnosed with DS-TB patients actually had MDR-TB at baseline, and 25/268 (9%) patients treated for DR-TB did not have rifampicin resistance detected on WGS. This may have been due previous mixed infection with other strains not found in the study sample, false positive/negative DST results or lab error.

All patients with DS-TB were treated with the standard treatment regimen (rifampicin, isoniazid, pyrazinamide and ethambutol). In line with global developments, there were substantial changes in DR-TB treatment regimens over the course of this study (2016-2019). At the beginning of this period, long regimens (20 months+) based on a fluoroquinolone and kanamycin were standard. For MDR-TB, these were progressively replaced with the short (nine-month) regimen based on kanamycin and a fluoroquinolone, and later bedaquiline and a fluoroquinolone. Pre-XDR/XDR-TB treatment remained long individualised regimens, although use of bedaquiline and linezolid became more common. Among the 268 patients with DR-TB for whom we have treatment data, all except three had been treated with bedaquiline or kanamycin: 128 received bedaquiline, 118 kanamycin and 19 received both or switched from kanamycin to bedaquiline. All except two patients with DR-TB received a fluoroquinolone. Full baseline treatment information in included in the Supplementary Spreadsheet.

There were 352 patients with sequences meeting the minimum coverage threshold of 80x included in the analysis of baseline isolates (collected at initiation or major change to treatment). We excluded 21/352 isolates (6.0%) that we characterised as mixed infection, and conducted detailed analyses on the remaining 331 single strain infections. We did not identify any patient or bacterial factors significantly associated with likelihood of mixed infection (Supplementary Table 2). Patients were only included in the analysis of sequential isolates if they had >1 successfully sequenced positive culture after excluding isolates with evidence of reinfection or superinfection since baseline (<15 SNP differences from baseline isolate) or mixed infection. There were 348 individual isolates from 118 patients with >1 timepoint at coverage above 55x included in the sequential analysis.

### Whole genome nucleotide diversity at baseline

Among single strain infections, there was no difference in median π when isolates were stratified by the number of PCR cycles (p=0.72, equality of medians test, Supplementary Figure 1). There was also no evidence for an association between π and the proportion of reads assigned to *Mycobacteria* (R-squared 0.6%, p=0.16 for linear association, Supplementary Figure 2), suggesting that high stringency mapping with BBMap adequately prevented mapping of contaminating reads. Finally, we tested whether isolates grown on solid media or after a single subculture in 7H9 were less diverse than those sequenced from MGIT culture, and again found no evidence for a statistically significant difference in π between groups (Supplementary Figure 3). We therefore concluded that the number of PCR cycles, isolate contamination and culture media did not significantly impact on within-host genetic diversity for the purposes of this study. As expected, mixed infections had significantly higher median π than single strain infections (7.75×10^−6^ v 9.48×10^−7^, p<0.001, Wilcoxon rank-sum test).

As bacterial load was expected to affect π, we tested for association with two measures of bacterial load: sputum smear grading and time to positivity of the original MGIT isolate (TTP). Smear grading is expected to increase and TTP to decrease with higher bacterial burden. There was no linear trend between increasing smear grade and π (R-squared 0.3%, p=0.43, Supplementary Figure 4), and while there was evidence for a linear trend of increase of π with higher TTP (coefficient 1.22×10^−8^, p=0.017, Supplementary Figure 5), it explained only a small proportion of the variance (R-squared 1.8%).

To investigate how π varied in patients starting/changing TB treatment, we tested its association with the following patient factors: age, sex, HIV status, CD4 count <200 (and taking antiretroviral therapy (ART)), cavitation on chest radiography, previous TB infection and if the patient was on TB treatment at the time of enrolment (for example for DS-TB prior to detection of drug resistance) (Table 2). We found that absence of cavitation on chest radiography (p=0.017, Wilcoxon rank-sum), taking ART (if HIV positive) (p=0.017) and who were taking TB treatment (p=0.049) at the time of enrolment were independently associated with a lower π on univariate analysis. In addition to these patient factors, we also examined bacterial lineage and drug resistance profile (Table 2). Lineages 2 and 4 are the major groups circulating in South Africa(49) which was also the case in our dataset. Patients infected with lineage 2 strains had greater π than those with lineage 4 infections (p<0.001). While there was no difference in π between patients with genetically DS-TB and TB genetically resistant to first line drugs (rifampicin/multidrug-resistant TB, RR/MDR-TB), patients with genetic second-line drug resistance (pre-/extensively drug-resistant TB, pre-XDR/XDR-TB) had lower π than patients with DS-TB (p<0.001) and RR-TB (p<0.001).

**Table 2.** Clinical and bacterial correlates of nucleotide diversity (π) in the 331 single strain baseline isolates downsampled to 80x mean coverage, showing number in each category, median π (× 10^−6^) and interquartile range, and p-value (Wilcoxon rank sum, except k-sample equality of medians where denoted by *).

| Characteristic | Number | Median $\pi \times 10^{-6}$<br>(IQR) | p value |
| --- | --- | --- | --- |
| **PATIENT FACTORS** |  |  |  |
| **Age category** |  |  |  |
| <25 | 32 | 0.995 (0.490-1.54) | 0.73* |
| 25-30 | 63 | 1.09 (0.619-1.48) |  |
| 30-35 | 63 | 0.981 (0.659-1.55) |  |
| 35-40 | 50 | 0.846 (0.489-1.15) |  |
| 40-50 | 72 | 0.961 (0.664-1.52) |  |
| $\geq 50$ | 45 | 0.863 (0.578-1.23) | |
| **Missing** | 6 |  |  |
| **Sex** |  |  |  |
| Male | 188 | 0.969 (0.640-1.55) | 0.31 |
| Female | 137 | 0.944 (0.563-1.39) |  |
| **Missing** | 6 |  |  |
| **Previous TB** |  |  |  |
| No | 167 | 0.973 (0.619-1.53) | 0.50 |
| Yes | 155 | 0.916 (0.578-1.41) |  |
| **Missing** | 9 |  |  |
| **HIV status** |  |  |  |
| Negative | 60 | 1.02 (0.678-1.48) | 0.33 |
| Positive | 268 | 0.939 (0.573-1.47) |  |
| **Missing** | 3 |  |  |
| **CD4 count (if HIV positive)** |  |  |  |
|  | 89 | 0.916 (0.587-1.33) | 0.41 |
| <200 | 106 | 0.831 (0.498-1.30) |  |
| $\geq 200$ | 73 | | |
| **Missing** |  |  |  |
| **Taking ART (if HIV positive)** |  |  |  |
| No | 66 | 1.16 (0.744-1.71) | 0.010 |
| Yes | 176 | 0.837 (0.562-1.34) |  |
| **Missing** | 26 |  |  |
| **Cavitation on chest radiography** |  |  |  |
| No | 89 | 0.725 (0.467-1.09) | 0.017 |
| Yes | 26 | 1.05 (0.668-1.59) |  |
| Missing | 216 |  |  |
| **On TB treatment at enrolment** |  |  |  |
| No | 153 | 1.03 (0.672-1.63) | 0.049 |
| Yes | 170 | 0.847 (0.563-1.36) |  |
| Missing | 8 |  |  |
| **BACTERIAL FACTORS** |  |  |  |
| **Mtb Lineage** |  |  |  |
| 1 | 10 | 1.49 (1.14-2.10) | Lineage 2 v 4<br><0.001 |
| 2 | 117 | 1.46 (1.02-1.93) |  |
| 3 | 8 | 1.24 (1.09-1.62) |  |
| 4 | 196 | 0.691 (0.486-1.04) |  |
| **Genetic drug resistance profile** |  |  |  |
| DS | 83 | 1.14 (0.668-1.71) | DS v MDR 0.22 |
| MDR | 179 | 1.03 (0.651-1.51) | DS v Pre-XDR/XDR <0.001 |
| Pre-XDR/XDR | 69 | 0.689 (0.446-0.948) | MDR v Pre-XDR/XDR <0.001 |

### Association between nucleotide diversity and outcome

We next tested if there was a correlation between baseline (initiation or major change to treatment) π and six-month outcome. Six-month outcomes were available for 297/331 (89.7%) patients with single strain infection, of whom 236/297 (79.5%) had a favourable six-month outcome. Unfavourable outcomes were due to death (24/297, 8.1%), loss to follow-up (20/297, 6.7%), and microbiological failure without acquired genetic resistance (11/297, 3.7%) or with acquired genetic drug resistance (6/297, 2.0%). We found that male sex, HIV coinfection and not taking ART therapy if HIV positive were associated with higher risk of unfavourable outcome (Supplementary Table 3). Interestingly, genetically MDR-TB infection was not associated with worse outcomes at month six than genetically DS-TB, although pre-XDR/XDR-TB infection was on univariate analysis (Table 2). Mixed infection, lineage, age, cavitation and previous TB infection also did not affect outcome. On univariate analysis, π was not associated with six-month outcome (Table 3). Not taking ART if HIV positive, pre-XDR/XDR-TB and taking TB treatment at the time of diagnosis (e.g. for DS-TB prior to an MDR-TB diagnosis) were considered as potential confounders and adjusted odds ratios were calculated. A final multivariate model was constructed including these potential confounders for both all patients (not including ART as a confounder) as well as HIV positive patients only (including ART as a confounder), but neither demonstrated an association between baseline π and six-month outcome (Table 3).

**Table 3.**
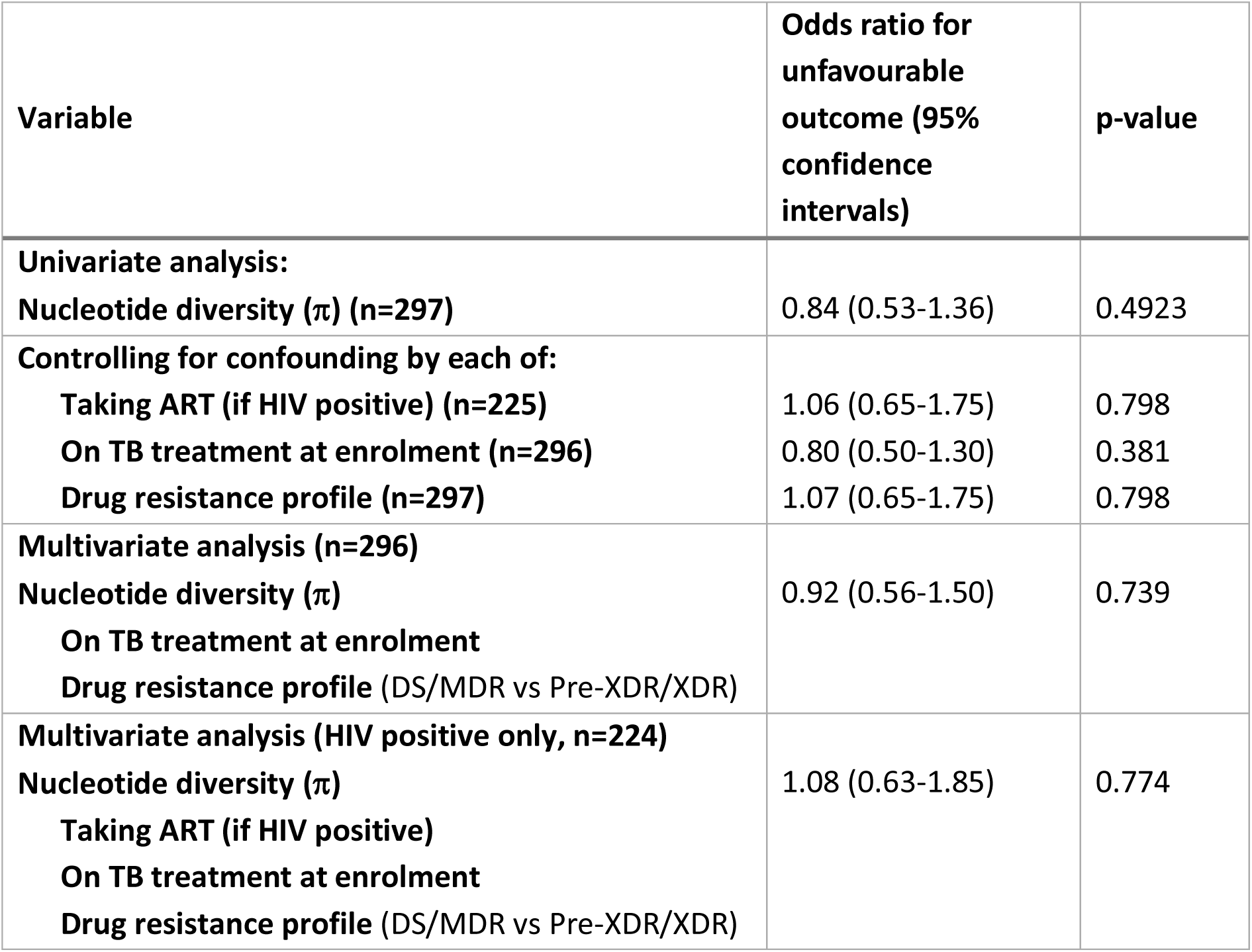
Univariate and multivariate logistic regression analyses for the association between baseline nucleotide diversity (π) and month six outcome.

### Dynamics of nucleotide diversity at the whole genome level during treatment

For the analysis of sequential isolates, all sequences (including baseline sequences) were downsampled to mean 55x genome-wide coverage. There were 118 patients included in this analysis, of which 115 had baseline isolates that met inclusion criteria. Three patients were excluded as they did not have a baseline sequence meeting criteria. Median π in these 115 isolates was 8.67×10^−7^, and not significantly different (p=0.104, Wilcoxon rank-sum test) to the value of 9.48×10^−7^ from the 331 baseline isolates downsampled to 80x analysed above. When comparing median π among all isolates at each timepoint, week 1 and 2 isolates showed an increase in π to 1.14×10^−6^ and 1.15×10^−6^ respectively (Figure 1), while there was no difference between baseline π and that at later timepoints. As this could be due to patients with less diverse baseline sequences culture converting rapidly, the analysis was repeated by including only patients who had a positive culture after week 2 of treatment, which yielded similar results (Supplementary Figure 6). The increase in π at weeks one and two remained statistically significant after correcting for multiple tests for both of these analyses. It is important to note that 61/115 (53%) of these patients were receiving TB treatment at the time of the baseline sample that was deemed to be ineffective due to programmatically-identified drug resistance, as is commonly the case for patients starting DR-TB treatment in South Africa. These patients had generally been taking treatment for one or more months and had by definition remained culture positive. However, it is possible that the changes in within-isolate diversity profiles for these patients over time may be different to those who were completely drug-naïve. When considering only the 54 patients who were not taking TB treatment at the time of enrolment, there were no statistically significant differences in π during treatment (Supplementary Figure 7).

**Figure 1.**
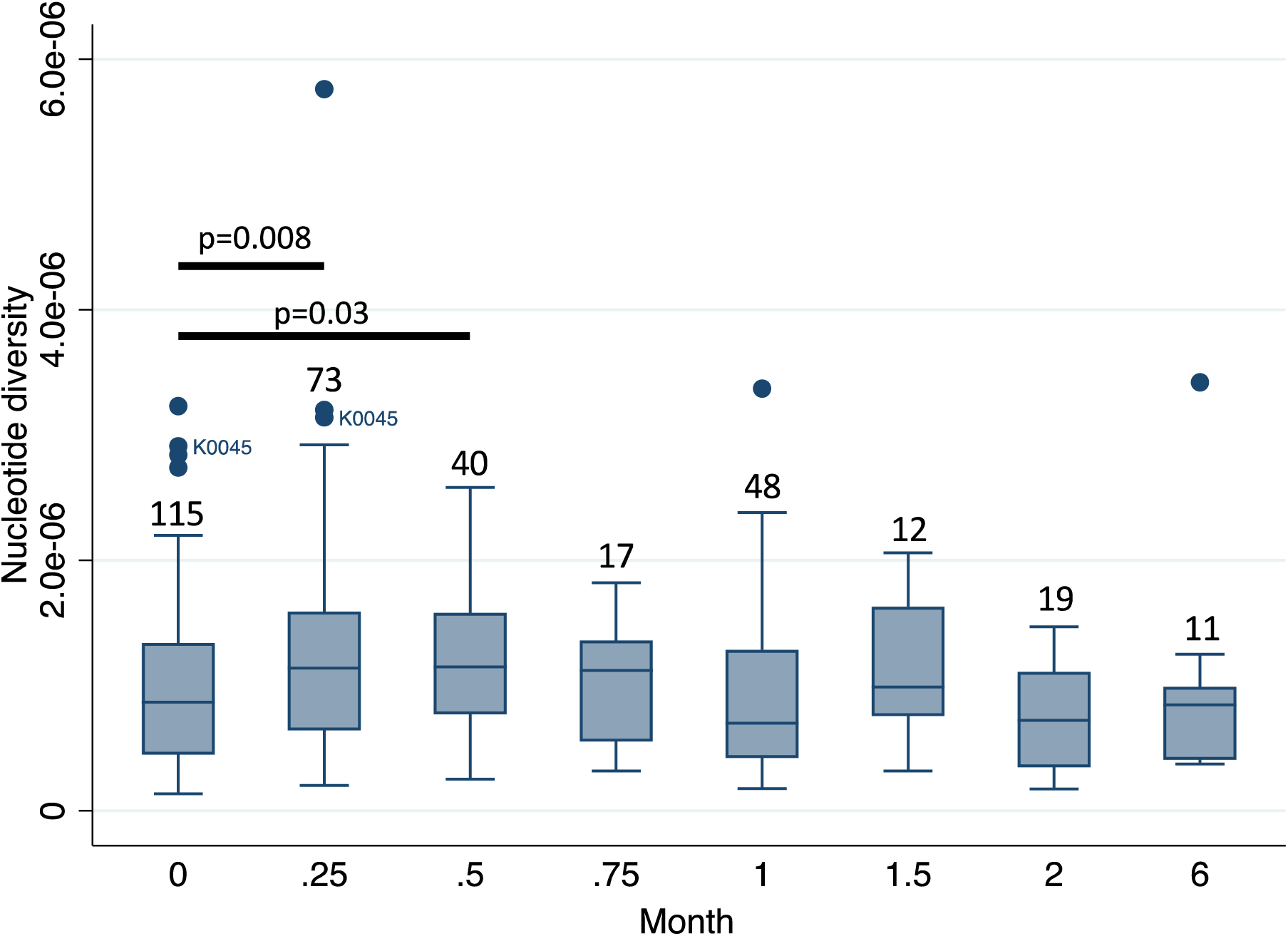
Box plot showing median and interquartile range of nucleotide diversity (π) in sequential isolates for timepoints where there were ≥5 isolates. Numbers indicate number of isolates at each timepoint. Circles mark outlying values and all represent different patients, except K0045 who was an outlier at baseline (month 0) and week 1 (month 0.25) and is labelled. Bars indicate where π statistically significantly differs from baseline (Wilcoxon rank-sum test).

### Individual gene-level nucleotide diversity

We then assessed within-isolate genetic diversity for all coding genes at baseline (initiation or major change to treatment). In addition to functional categories, two additional redundant categories of antibiotic resistance genes and antigen genes were generated as described in the methods. There was no difference in mean π between any gene or functional category in the 331 single strain baseline isolates downsampled to 80x coverage (Table 4). The most diverse coding genes at baseline are listed in Supplementary Table 4, although many are not yet well described. In line with previous results(15), *Rv1319c*, a putative adenylate cyclase responsible for regulation of cellular metabolism was the gene displaying the highest within-host genetic diversity.

**Table 4.**
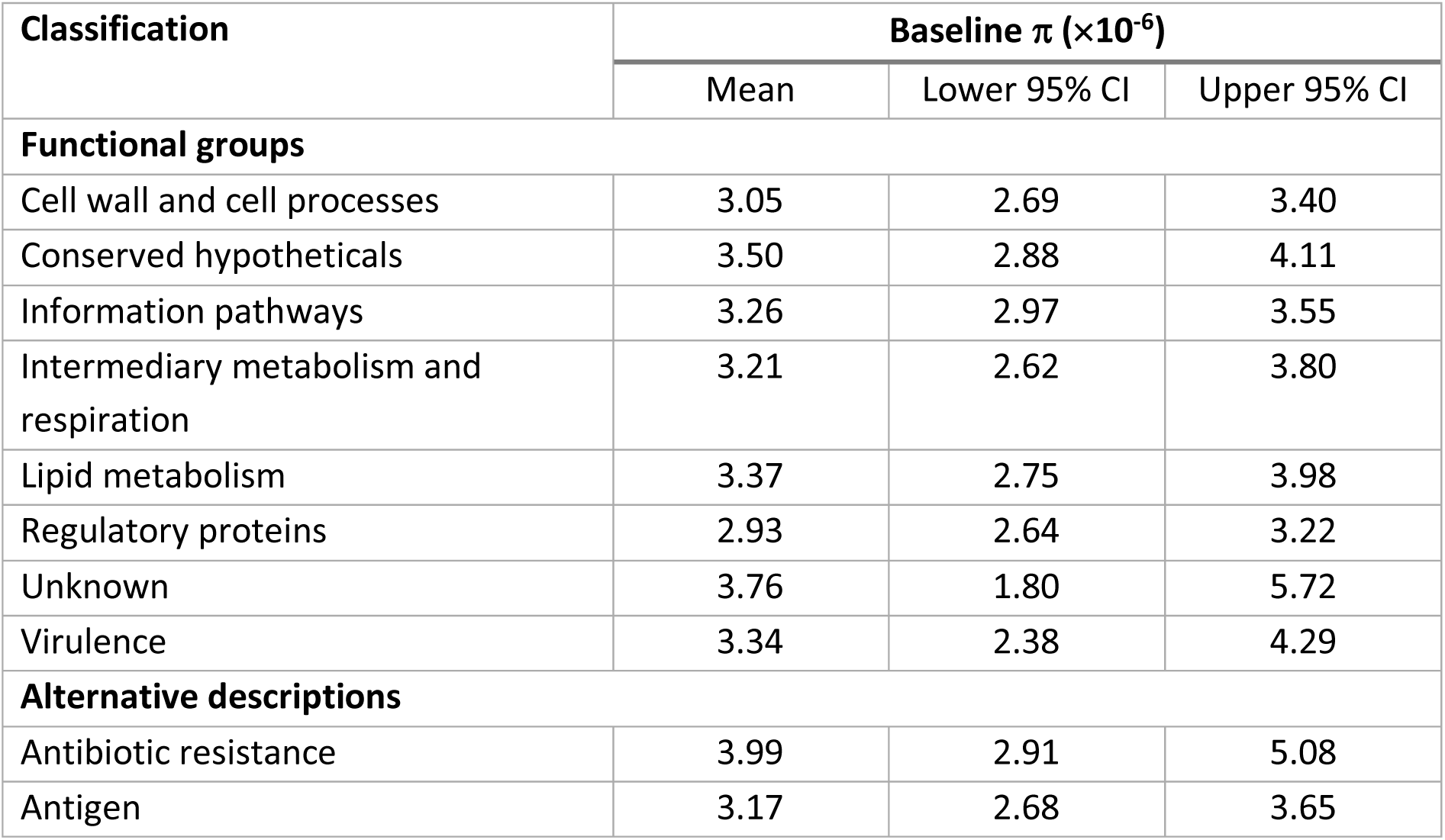
Mean nucleotide diversity (π) and 95% confidence intervals (expressed at ×10^−6^) in baseline isolates by functional group and gene description.

| Classification | Baseline $\pi$ ($\times 10^{-6}$) | | |
| --- | --- | --- | --- |
|  | Mean | Lower 95% CI | Upper 95% CI |
| **Functional groups** |  |  |  |
| Cell wall and cell processes | 3.05 | 2.69 | 3.40 |
| Conserved hypotheticals | 3.50 | 2.88 | 4.11 |
| Information pathways | 3.26 | 2.97 | 3.55 |
| Intermediary metabolism and respiration | 3.21 | 2.62 | 3.80 |
| Lipid metabolism | 3.37 | 2.75 | 3.98 |
| Regulatory proteins | 2.93 | 2.64 | 3.22 |
| Unknown | 3.76 | 1.80 | 5.72 |
| Virulence | 3.34 | 2.38 | 4.29 |
| **Alternative descriptions** |  |  |  |
| Antibiotic resistance | 3.99 | 2.91 | 5.08 |
| Antigen | 3.17 | 2.68 | 3.65 |

To establish which functional categories of genes were driving changes in π over time, we examined π in the sequential isolates from the 115 patients described above. Diversity increased across multiple functional categories at the week 1 timepoint, before decreasing but remaining more diverse than at baseline (Figure 2**Error! Reference source not found**.). At each timepoint different genes were responsible for the changes in diversity seen (Table 5). The changes in mean π were seen at month six are likely influenced by the small number of patients remaining culture positive (11 patients). However there was a notable increase in antibiotic resistance gene π at this timepoint, driven by hetRAVs in *Rv0678*, a gene responsible for bedaquiline resistance, and which occurred exclusively in patients who were treated with bedaquiline and subsequently acquired clinical resistance.

**Table 5.** Genes with greatest increase in nucleotide diversity (π) (expressed at ×10^−6^) between timepoints.

| Order | Baseline → Week 1 | | Baseline → Month 2 | | Baseline → Month 6 | |
| --- | --- | --- | --- | --- | --- | --- |
| | Gene | $\Delta\pi (\times 10^{-6})$ | Gene | $\Delta\pi (\times 10^{-6})$ | Gene | $\Delta\pi (\times 10^{-6})$ |
| 1 | *Rv1319c* | 62.7 | *accE5* | 263.2 | *Rv0678* | 302.7 |
| 2 | *Rv2319c* | 58.4 | *Rv1765c* | 117.1 | *cysA2* | 136.3 |
| 3 | *Rv3424c* | 55.7 | *Rv2876* | 111.5 | *Rv3612c* | 113.4 |
| 4 | *Rv0219* | 54.3 | *cfp6* | 105.9 | *Rv2980* | 124.9 |
| 5 | *Rv1083* | 53.8 | *Rv1498c* | 73.4 | *Rv3655c* | 123.6 |
| 6 | *Rv1435c* | 51.6 | *cysA2* | 72.3 | *Rv0100* | 119.6 |
| 7 | *moaD1* | 48.8 | *mmpS4* | 70.8 | *Rv1948c* | 118.1 |
| 8 | *mscL* | 44.1 | *parE1* | 69.3 | *Rv2081c* | 92.0 |
| 9 | *Rv0634A* | 44.1 | *serB1* | 57.8 | *Rv1134* | 89.3 |
| 10 | *Rv0579* | 36.8 | *Rv3412* | 55.3 | *Rv1000c* | 80.9 |

**Figure 2.**
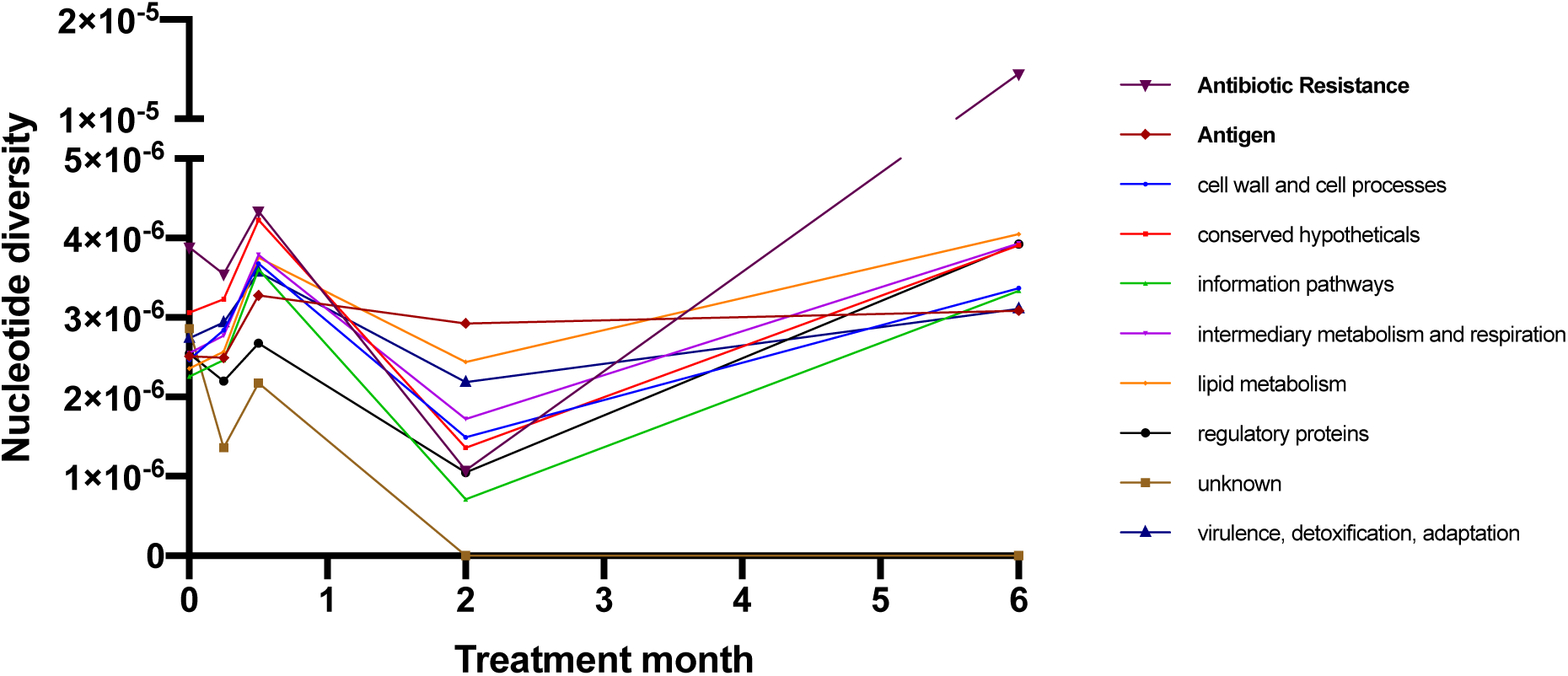
Change in mean nucleotide diversity (π) over time according to gene functional categories (solid icons). Two additional redundant categories are also plotted (bold text).

### Heteroresistance

We then assessed the prevalence of heteroresistance in isolates with a minimum coverage depth of 55x that were not mixed infections. There were 399 baseline isolates included in this analysis, with genetic resistance (presence of ≥1 heterozygous or fixed RAV) most frequently identified in baseline isolates for rifampicin (77.1%) and high-level isoniazid resistance (*katG* RAVs, 60.4%) as shown in Figure 3a in keeping with the sampling strategy for this study. Genetic resistance conferred only by hetRAVs was proportionately most common for bedaquiline, where the majority (60.0%, 6/10) of isolates contained only heterozygous RAVs, and fluoroquinolones (12.7%, 9/62) as shown in Figure 3b. Examining all baseline hetRAVs (including where multiple hetRAVs occurred in one isolate or where they were combined with fixed RAVs), allele frequencies varied from the limit of detection (7% at the minimum coverage of 55x, or 3% at coverage of 150x) to our nominated cut-off for fixation of 95% (Figure 3c).

**Figure 3.**
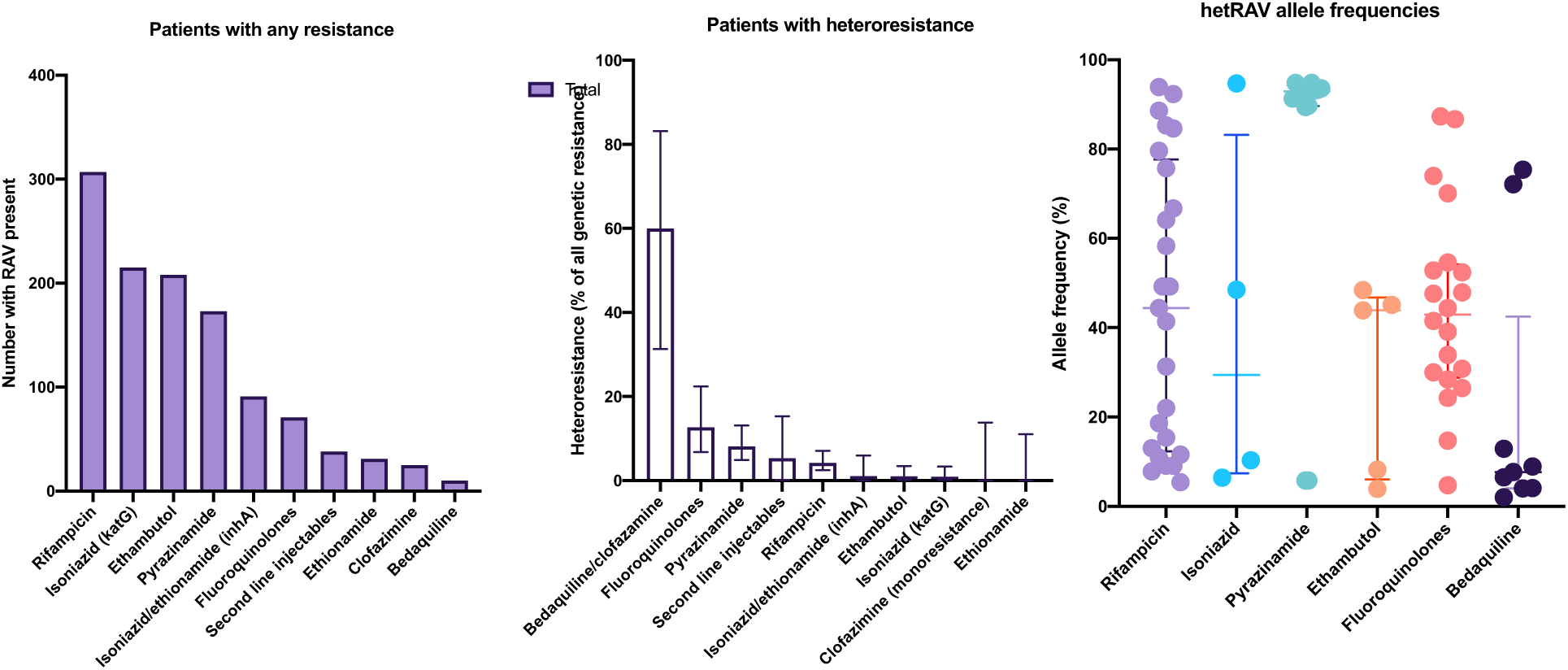
(a) Number of baseline isolates with genetic resistance (i.e. containing any RAV) by drug (b) percentage of baseline isolates with hetRAVs only amongst all RAVs and (c) baseline allele frequency of all hetRAVs by drug, with lines showing median and interquartile range. Some isolates have >1 hetRAV.

Where longitudinal sampling was available, the majority of patients with resistance conferred by hetRAVs had either one resistance mutation reaching fixation (9/20, 45%) or heteroresistance persisting with at least one hetRAV (8/20, 40%) (Table 6, full index of all hetRAVs in Supplementary Table 5). In three cases (15%) hetRAVs disappeared altogether: one *Rv0678* M146T mutation at 2.0% frequency (conferring bedaquiline resistance), one *rpoB* S450L mutation at 9.1% frequency (rifampicin resistance) and one *embB* D328Y mutation at 43.9% frequency (ethambutol resistance). In the five cases of rifampicin heteroresistance that became fixed or persisted, all patients actually had two separate hetRAVs, all of which displayed dynamics consistent with clonal interference of two resistant subpopulations (Figure 4a-e). Rifampicin had already been stopped in all five patients following diagnosis of MDR-TB. Clonal interference was also seen in both patients with fluoroquinolone heteroresistance (Figure 4f-g) and one of the patients with bedaquiline/clofazimine heteroresistance (Figure 4h), and all of these patients were still treated with the respective drugs. All pyrazinamide heteroresistance was caused by T153fs mutations at high frequency, that were at or close to the arbitrary cut-off of 95% for fixation.

**Table 6.** Resistance patterns in longitudinal isolates from patients with resistance conferred only by heterozygous resistance-associated variants (hetRAVs) at baseline. Numbers refer to patients.

| **Drug** | **Patients with hetRAV presence in further isolate(s) (%)** |  |  |  |
| --- | --- | --- | --- | --- |
|  | **Total number** | **RAV(s) become fixed** | **RAV(s) remain heterozygous** | **RAV(s) disappear** |
| Rifampicin | 6 | 4 (66.7%) | 1 (16.7%) | 1 (16.7%) |
| Isoniazid | 1 | - | 1 (100.0%) | - |
| Pyrazinamide | 6 | 3 (50.0%) | 3 (50.0%) | - |
| Ethambutol | 1 | - | - | 1 (100.0%) |
| Fluoroquinolones | 2 | 1 (50.0%) | 1 (50.0%) | - |
| Second line Injectables | 1 | - | 1 (100.0%) | - |
| Bedaquiline | 3 | 1 (33.3%) | 1 (33.3%) | 1 (33.3%) |

**Figure 4.**
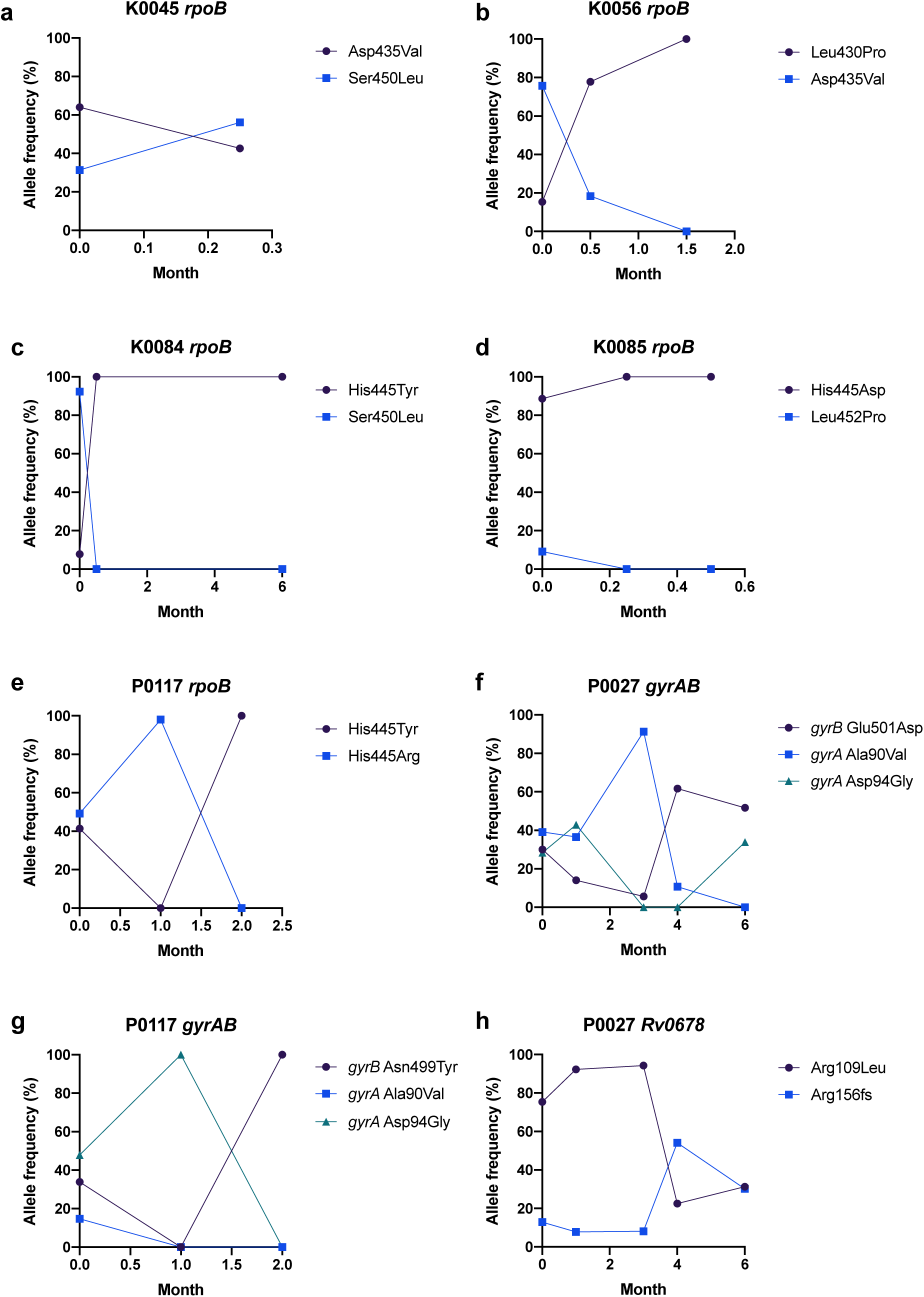
Allele frequency graphs for patients with evidence of clonal interference in *rpoB* (a-e), *gyrAB* (f-g) and *Rv0678* (h).

### Predicting emergent resistance

Emergent resistance in this cohort was rare, with only 9/286 (3.1%) of patients from cohorts where follow-up samples were routinely collected (studies one to four in Table 1) identified as acquiring RAVs after exclusion of mixed infections and reinfections. Among the 77 patients with fully drug-susceptible TB (DS-TB) one patient acquired a rifampicin RAV when they relapsed at month 12, having initially culture converted at week 3, and one patient acquired an isoniazid RAV. Additionally, one patient acquired a fluoroquinolone RAV, and six patients acquired bedaquiline and clofazimine cross-resistance RAVs, one of whom also acquired delamanid RAVs (Table 7). All new RAVs occurred in the presence of treatment with the drug. All bedaquiline resistance was conferred by RAVs in the efflux pump repressor gene *Rv0678*, with no RAVs identified in the other resistance-associated genes *atpE* or *pepQ*, which have been more frequently associated with resistance *in vitro* or murine model studies(50,51).

**Table 7.** Emergent RAVs occurring while on treatment in patients who were previously wild-type in the context of single strain infections. The month of RAV emergence as identified by WGS is shown and the frequency at this time, followed by results of retrospective targeted deep sequencing of any isolates preceding RAV emergence on WGS. Dashes indicate sequencing data not available for these isolates. *this patient was culture negative after six months’ treatment for isoniazid monoresistant TB, but relapsed month 12 with new rifampicin resistance.

| Patient ID | Drug | Mutation | Month of emergence | Frequency at emergence (WGS) | Frequency in retrospective deep sequencing of isolate preceding emergence |
| --- | --- | --- | --- | --- | --- |
| R0075 | Rifampicin | *rpoB* D435V<br>*rpoB* S450L | 12*<br>12* | 83.3%<br>17.2% | absent<br>absent |
| K0064 | Isoniazid | *katG* W328stop | 0.25 | 5.9% | - |
| P0064 | Fluoroquinolones | *gyrA* D94N | 2 | 47.6% | absent |
| K0061 | Bedaquiline | *Rv0678* G138stop | 0.25 | 4.5% | - |
| P0082 | Bedaquiline | *Rv0678* C46fs<br>*Rv0678* D47fs | 5<br>4 | 92.6%<br>55.6% | absent<br>absent |
| P0200 | Bedaquiline | *Rv0678* D47fs | 6 | 95.3% | absent |
| P3505 | Bedaquiline | *Rv0678* G147fs | 6 | 94.5% | absent |
| P3524 | Bedaquiline | *Rv0678* Q22P | 2 | 85.1% | absent |
|  |  | *Rv0678* D47fs | 2 | 45.2% | absent |
|  |  | *Rv0678* A57E | 6 | 10.6% | 0.55% |
|  |  | *Rv0678* R72T | 6 | 9.2% | absent |
|  |  | *Rv0678* D88fs | 6 | 12.0% | absent |
|  |  | *Rv0678* D88A | 6 | 13.0% | absent |
|  |  | *Rv0678* G121R | 6 | 6.0% | 1.09% |
|  |  | *Rv0678* L122P | 6 | 7.4% | 0.94% |
|  | Delamanid | *fbiC* A487fs<br>*fbiC* S534stop | 6<br>6 | 24.7%<br>11.9% | absent<br>absent |
| K0084 | Bedaquiline | *Rv0678* V1A | 6 | 60.0% | absent |
|  |  | *Rv0678* G37fs | 6 | 11.9% | absent |
|  |  | *Rv0678* R89W | 6 | 12.5% | absent |

To establish if emergence of genetic resistance could be predicted by presence of low-frequency (≤1%) variants in preceding isolates, we performed deep sequencing of resistance-associated genes (Supplementary Table 1) with minimum and mean coverage of 2897x and 4072x respectively for seven of the nine patients with emergent resistance and sufficient surplus DNA after standard WGS. Deep sequencing revealed large numbers of low-frequency variants. For example, the mean number of *Rv0678* variants called was 66 over all baseline isolates. Despite this, none of the emergent RAVs described in Table 7 were identified in the preceding deep-sequenced isolates and no phenotypic resistance was identified to any drug tested prior to the emergence of a RAV on WGS, with the exception of participant P3524 where three of five additional bedaquiline/clofazimine RAVs that appeared at month six were present at frequencies around 1% in the month 2 isolate. The month 2 isolate was phenotypically bedaquiline and clofazimine resistant but also had bedaquiline RAVs at >5% frequency. Further use of deep sequencing to identify low-frequency variants will require a system validated to detect a range of allele mixtures.

## Discussion

In this study we have analysed within-host diversity of *Mtb* isolates at the whole genome level, across gene classes and at specific resistance-associated sites in several South African TB cohorts. A particular strength of this study is that all cohorts were prospective, reducing the potential for bias in patient selection and by limiting the number of subculturing steps prior to and following isolate storage.

The 6% rate of mixed infection that we identified is not as high as the 10-20% that has been reported in other studies in high-burden settings(9,52), especially considering that WGS offers additional sensitivity to detect mixed infections compared to previous technologies. This may be because time to treatment initiation has decreased with health systems changes, including introduction of the Xpert MTB/RIF rapid diagnostic(53,54). We did not identify any factors associated with risk of mixed infection and contrary to previous studies did not detect worse outcomes in patients with mixed infection, although we compared six-month rather than end of treatment outcome and our studies were heavily weighted for patients with DR-TB.

We sought to determine if technical factors such as subculturing in liquid or solid media and isolate contamination could affect within-host genetic diversity, and reassuringly none had a measurable effect. While subculturing has been shown to lead to a loss in genetic diversity, we limited subculturing to a single event, tried to transfer the maximum volume of liquid culture possible or scrape all colonies from solid plates. To prevent cross-mapping of contaminating reads artefactually inflating within-host genetic diversity, we used sequence read mapping software that allows stringent read-identity parameters to be set which appeared effective on this dataset.

We expected diversity to be related to bacterial load in the original patient sample but the absence of a correlation with smear positivity or TTP may be because these are not ideal measures of true within-patient bacterial load, being inherently limited by the stochasticity of sputum sampling and potential culture-induced bias. Conversely other factors such as drug penetration or immune control may be more important drivers of within-host genetic diversity than bacterial population size. Supporting this, we did find greater genetic diversity in patients with cavitary disease which may lead to spatially separated bacterial subpopulations, likely exposed to a different microenvironment for example in respect to drug concentrations (Table 2). We also observed higher within-host genetic diversity in patients who were HIV positive but not taking ART suggesting an effect of immune constitution not captured by CD4 count. *Mtb* lineage 2 was correlated with greater diversity than lineage 4, supportive of the proposed higher mutation rate in lineage 2(55,56). DS and MDR-TB infections had similar genetic diversity, while pre-XDR/XDR were less diverse and may represent the likely fitness costs of these additional mutations(57) (Table 2).

Overall, clinical outcomes were excellent with similar six-month outcomes in DS and MDR-TB, backing up recent promising observational reports from MDR-TB programmes(58), although six-month outcomes may not fully translate into end of treatment outcomes(59). While mixed infections have previously been reported to be associated with poor end of treatment outcomes(5,6), in this study baseline nucleotide diversity (π) at the point of initiating or undergoing a major change in treatment regimen in single strain infection was not associated with six-month outcome (Table 3). Although we did not identify worse outcomes in patients with mixed infection (which may be as numbers were small), it suggests that the association between mixed infection and poor outcomes reported elsewhere(5,6) is mediated by a process independent of genomic diversity.

There was a significant increase in overall genomic π at weeks 1-2, which then reduced back to similar levels to baseline throughout the rest of treatment (Figure 1). There was no evidence of a decrease from baseline in π during the later stages of treatment, although the number of patients who were culture positive at later timepoints was small, and inevitably successfully treated patients become culture negative within the first weeks of treatment. The initial increase in diversity was driven by increases in π of multiple different gene functional categories (Figure 2). While diversity appeared to increase again at month six, numbers were small at this timepoint. As shown previously, we observed no enrichment of T cell antigens or antibiotic resistance genes(14), except for the increase in diversity in antibiotic resistance genes at month six driven by hetRAVs in *Rv0678* among patients who had acquired bedaquiline resistance.

Frequency of genetic heteroresistance varied considerably by drug and was most common for bedaquiline and fluoroquinolones and rare for isoniazid and rifampicin in line with previous descriptions(44) (Figure 3b). In contrast to other drugs, pyrazinamide heteroresistance was largely caused by a single T153fs mutation at >90% frequency that did not reach fixation, potentially as the associated fitness cost(60) causes an evolutionary pressure to maintain the wild type allele (although the fitness implications of *pncA* mutations remain disputed(61)). *Rv0678* bedaquiline/clofazimine RAVs often persisted as heterozygous variants often throughout treatment, suggesting there is an as yet undetermined underlying biological explanation why they fail to reach fixation. A driving factor behind heteroresistance to other antimycobacterial drugs may be the existence of *Mtb* subpopulations in lung cavities, where some drugs may penetrate at subtherapeutic concentrations or not at all(20,62). While this has not yet been conclusively demonstrated for bedaquiline, along with clofazimine it is predicted to exhibit poor penetration into cavities(63).

While the majority of hetRAVs persisted or became fixed, some of those at lower frequencies disappeared in sequential sampling. The current allele frequency limit of detection of Xpert MTB/RIF for rifampicin heteroresistance by Xpert MTB/RIF is 20-80%(27) and 5% for fluoroquinolone heteroresistance by line probe assays(28). It is important that future rapid molecular diagnostics and whole genome sequencing pipelines are designed to identify and quantify genetic heteroresistance, as allele frequency may affect the clinical significance of these mutations. Most patients with rifampicin and all with fluoroquinolone heteroresistance demonstrated fluctuating hetRAV frequencies consistent with clonal interference (Figure 4), suggesting the presence of more than one subclone co-existing for weeks or months within a patient, rather than single dominant clone.

Emergent resistance during treatment was rare in this study compared to reports from only a few years ago quoting acquired second-line drug resistance in >25% of patients with MDR-TB during treatment(64). This could be due to the now widespread use of bedaquiline and other bactericidal drugs which have been shown to reduce acquisition of second line drug resistance(65) and other programmatic changes including introduction of rapid molecular diagnostics for resistance and the nine-month regimen in addition to the rigorous exclusion of mixed and reinfection by WGS of all isolates. *Rv0678* variants conferring bedaquiline/clofazimine cross-resistance accounted for the majority of emergent resistance mutations, which is likely to reflect both the relative importance of these drugs to current regimens and also that it may be comparatively easy for *Mtb* to acquire resistance due to the large mutational target for bedaquiline/clofazimine resistance with any mutation disrupting the function of the *Rv0678* gene removing repression of the MmpL5 efflux pump.

While there have been several reports of microheteroresistance (RAVs with <5% allele frequency) RAVs pre-dating the emergence of macroheteroresistance (RAVs with 5-95% allele frequency) or fixed resistance, in this prospectively enrolled cohort we found a high degree of noise with large numbers of very low-frequency variants detected on retrospective deep sequencing of isolates collected preceding the emergence of resistance of WGS. This may reflect either high turnover of variants that are rapidly selected against and/or sequencing error, and effectively separating the two remains challenging without a gold standard to discriminate the two. SureSelect RNA bait enrichment has previously been shown to not to bias population structure of heterozygous alleles with frequency >1%(66), but should be studied further in *Mtb* using a culture and sequencing pipeline that has been formally validated with *in vitro* allele mixtures of different frequencies covering the range of interest. However, as very few of the emergent RAVs were identified even among the multiple low-frequency variants that may represent standing variation or sequencing error(2), it suggests that weekly or more frequent sampling in addition to effective exclusion of sequencing error would be required to identify low-frequency RAVs that were likely to be clinically significant. This is in agreement with another study of treatment failures that found that only variants at >19% frequency were likely to become fixed(14) and that very low-frequency variants did not affect outcome of patients with DS-TB(5).

Limitations of this study include that, as for the majority of studies of *Mtb*, WGS was performed on cultured isolates that may not truly represent the true within-host diversity. Sequencing directly from sputum is now possible but has not been implemented on sufficient scale to utilise during this study period. Not all cohorts collected samples at all timepoints, with week 1 data sourced from two cohorts and week 2 data from one cohort. Some patients were already taking treatment at the time of study entry, and although to be enrolled the patient would have to still be culture positive and require addition of ≥2 new drugs, it is possible that pre-treatment may have affected genetic diversity. Additionally, we selected cohorts that primarily recruited patients with DR-TB as these patients were expected to be at higher risk of acquiring further drug resistance and have prolonged culture positivity compared to patients with DS-TB. Therefore, while the findings may be true for these cohorts which ultimately had good treatment outcomes and relatively rare acquisition of further resistance, they do not necessarily represent a true cross-section of patients with TB in South Africa or elsewhere in the world.

In conclusion, in this prospective study of South African patients starting TB treatment, most of whom had at least first line drug-resistance, there was no association between within-host genetic diversity and clinical outcome for patients infected with a single strain. Studying within-host *Mtb* genetic diversity identifies changes in genetic diversity during treatment and is a potential tool to investigate the bacterial response to selective pressures. Further work is required to delineate allele frequency thresholds for identification of significant low-frequency RAVs, and frequent sampling may be required to identify them. The emergence of bedaquiline/clofazimine and delamanid resistance is concerning and highlights the need for rigorous antimicrobial stewardship and resistance monitoring.

## Data Availability

Anonymised identifiers for patients included in all analyses and corresponding NCBI SRA accessions are listed in the Supplementary Spreadsheet. All whole genome sequencing data are available under the NCBI SRA BioProject PRJNA559528.

## Funding Sources

CN is supported by the Wellcome Trust (203583/Z/16/Z). JM is supported by the Wellcome Trust (203919/Z/16/Z). MO is supported by National Institutes of Health/National Institute of Allergy and Infectious Diseases (R01AI124413). FB acknowledges support from the Medical Research Council (MRC) (MR/P007597/1), the BBSRC (BB/R01356X/1) and the National Institute for Health Research University College London Hospitals Biomedical Research Centre. The funders did not have any role in study design, data collection, data analysis, interpretation, or writing of the report.

## Declaration of interests

Camus Nimmo and James Millard declare grants from the Wellcome Trust. Alexander S. Pym is now employed by Janssen Pharmaceutica. Max O’Donnell declares grants from NIH/NIAID. François Balloux declares grants from MRC, BBSRC and NIHR UCLH BRC.

## Author Contributions

Study design: CN, JB, FB

Data collection: CN, KB, JM, AG, NP, MO

Data analysis: CN, FB

Data interpretation: CN, JM, MO, FB

Writing: CN, FB

Review and approval of manuscript: CN, KB, JM, AG, NP, MO, JB, FB

All authors have read and approved the final version of this manuscript.

## Acknowledgements

The authors would like to thank Rachel Williams, Erika Yara Romero, Charlotte Williams, Helena Tutill and Patricia Dyal Bynoe from the University College London Pathogen Genomics Unit for assistance with whole genome sequencing.

## Supplementary Material

Supplementary Spreadsheet. List of isolates included in each analysis and accession numbers.

Supplementary Methods 1. Phenotypic drug susceptibility testing drugs and critical concentrations

Supplementary Methods 2. Method for DNA extraction

Supplementary Methods 3. Genetic mutations conferring drug resistance

Supplementary Table 1. Genomic regions covered by resistance bait set.

Supplementary Table 2. Associations between mixed infection and patient and bacterial factors.

Supplementary Table 3. Correlates of month six outcome showing hazard ratio and 95% confidence intervals (95% CI).

Supplementary Table 4. Classification, description and nucleotide diversity (π) of 10 most diverse coding genes in baseline sequences.

Supplementary Table 5. Frequencies and outcomes of baseline hetRAVs.

Supplementary Figure 1. Nucleotide diversity (π) median and interquartile range by number of PCR cycles in library preparation. Outlying values are marked with circles. There were no statistically significant differences between any of the distributions (Wilcoxon rank-sum test).

Supplementary Figure 2. Scatter plot and linear regression line of proportion of *k*-mers assigned to *Mycobacteria* by Kraken2 and nucleotide diversity.

Supplementary Figure 3. Nucleotide diversity (π) by isolate culture media: MGIT (initial culture), 7H9 (single subculture), 7H11 (solid plate) or unknown. There were no statistically significant differences between any of the distributions (Wilcoxon rank-sum test).

Supplementary Figure 4. Linear regression of increasing smear grade level against nucleotide diversity (π) with fitted regression line.

Supplementary Figure 5. Linear regression of increasing MGIT time to positivity (TTP) against nucleotide diversity (π) and fitted regression line.

Supplementary Figure 6. Box plot showing median and interquartile range of nucleotide diversity (π) in sequential isolates from patients who had at least one positive culture after week 2. Circles mark outlying values. Numbers indicate number of isolates at each timepoint. Bars indicate where distribution significantly different from baseline. *p=0.015 **p=0.006

Supplementary Figure 7. Box plot showing median and interquartile range of nucleotide diversity (π) in sequential isolates from patients who were not receiving any TB treatment at the time of enrolment.

